# Stratified Burden of MASLD in Cardiovascular-Kidney-Metabolic syndrome: Stage-Dependent Prevalence and Insulin Resistance-Driven Cardiovascular Mortality

**DOI:** 10.1101/2025.04.07.25325431

**Authors:** Zhuoxing Li, Qianyu Yang, Mao Xiao, Xue Zhang, Yanyi Deng, Hao Liu, Xiunan Liu, Yun Sun, Xiang Xiao

**Affiliations:** Department of Clinical Medicine, Chengdu Medical College, Chengdu, China; Department of Nephrology, The first affiliated hospital of Chengdu Medical College, China; Department of Nursing, Chengdu Medical College, Chengdu, China

**Author notes:** Correspondence: Yun Sun, MD, Email: 188487007 @qq.com, Department of Clinical Medicine, Chengdu Medical College., Address: No. 278, Middle Section of Baoguang Avenue, Xindu District, Chengdu, 610500, Sichuan, China., Xiang Xiao, PhD, Department of Nephrology, The first affiliated hospital of Chengdu Medical College. Address: No. 278, Middle Section of Baoguang Avenue, Xindu District, Chengdu, 610500, Sichuan, China. These authors have contributed equally to this work (Zhuoxing Li, Qianyu Yang, Xiunan Liu).

**Keywords:** metabolic dysfunction-associated steatotic liver disease, Cardiovascular-Kidney-Metabolic syndrome, epidemiological, insulin resistance, TyG index, Cardiovascular Mortality

## Abstract

**Objective:** This study aimed to investigate the epidemiological burden of metabolic dysfunction-associated steatotic liver disease (MASLD) across Cardiovascular-Kidney-Metabolic syndrome (CKM) stages and evaluate its association with cardiovascular mortality, while exploring the mediating role of insulin resistance (IR).

**Methods:** Using data from the National Health and Nutrition Examination Survey (NHANES, 2009–2018), we included 9,093 adults with CKM stages 1–4. MASLD was defined by validated indices (usFLI ≥ 30). Weighted Cox regression assessed MASLD-associated cardiovascular mortality risk. Restricted cubic splines (RCS) modeled dose-response relationships. Causal mediation analysis quantified TyG index’s contribution to MASLD-related mortality. Sensitivity analyses included subgroup stratification, missing data deleting and alternative MASLD definitions.

**Results:** MASLD prevalence increased significantly across advancing CKM stages (stage 1: 8.04%, stage 2:32.78%, stage 3: 41.90% and stage 4: 42.55%; *P* < 0.001). RCS revealed linear mortality risk escalation with rising usFLI scores (Non-line *P* < 0.05). MASLD independently predicted 63% higher cardiovascular mortality risk (adjusted HR=1.63, 95% CI:1.05–2.52). Stratify analyses revealed heterogeneity in associations by diabetes, CKD, CVD, and CKM stages (*P* for interaction < 0.05), stronger risks were observed in non-diabetic, non-CKD, non-CVD and early-stage (1-2) CKM. TyG-mediated IR explained 40.5% of MASLD-associated mortality. Sensitivity analyses confirmed robustness across MASLD definitions (FLI-based HR = 1.68, 95% CI, 1.07 - 2.63, *P* = 0.025).

**Conclusion:** MASLD exhibits a stage-dependent escalation in CKM populations and independently drives CVD mortality, with insulin resistance mediating 40% of this risk. Integrating MASLD screening into CKM risk stratification may enhance early intervention, particularly in early-stage patients.

## 1 Introduction

Metabolic dysfunction-associated steatotic liver disease (MASLD), previously termed nonalcoholic fatty liver disease (NAFLD), is the most prevalent chronic liver disease globally, affecting approximately 25% of adults worldwide. Its incidence has paralleled the rising prevalence of obesity and metabolic syndrome [1].Insulin resistance (IR) is the primary pathogenic driver of MASLD, promoting hepatic lipid deposition, chronic inflammation, and oxidative stress, which collectively contribute to multiorgan metabolic dysregulation involving the liver (steatosis to fibrosis), pancreas (β-cell dysfunction), vasculature (endothelial impairment), and adipose tissue (proinflammatory cytokine release) [2, 3].Furthermore, MASLD exacerbates IR, establishing a vicious cycle [4].This systemic metabolic dysfunction positions MASLD as an independent risk factor for type 2 diabetes mellitus (T2DM) and atherosclerotic cardiovascular disease (ASCVD) [5, 6].

Recently, the American Heart Association introduced the concept of cardiometabolic-kidney syndrome (CKM), emphasizing the interplay among cardiac, renal, and metabolic systems. The CKM staging system classifies disease progression into five phases (from risk factor accumulation to end-organ damage), with inflammation, oxidative stress, IR, and vascular dysfunction identified as core pathways driving metabolic risk progression, kidney disease evolution, enhanced cardiorenal interactions, and cardiovascular disease (CVD) development [7].However, current CKM staging primarily relies on diabetes, hypertension, and chronic kidney disease (CKD) for risk stratification, neglecting MASLD—a pivotal metabolic liver phenotype [8].This omission may underestimate the liver’s regulatory role in multiorgan metabolic axes. The liver serves not only as a primary target of IR but also secretes hepatokines (e.g., fibrinogen-like protein 2, fetuin-A) that directly promote vascular inflammation and renal injury [9–11]. Notably, MASLD exhibits high comorbidity with CKM components such as obesity, T2DM, and CKD [12].Global data synthesized by Younossi Z et al. [13] reveal that MASLD (diagnosed via ultrasound or MRI) coexists in 55–75% of T2DM patients, reaching 70.6% in Asian populations. Additionally, MASLD patients demonstrate a 1.2- fold increased prevalence of CKD and a 1.96-fold elevated risk of abnormal albuminuria compared to controls[14].

Nevertheless, large-scale clinical studies remain scarce regarding the epidemiological characteristics of MASLD in CKM populations and its impact on CVD outcomes, particularly MASLD’s contribution within the CKM framework. Moreover, synergistic mechanisms between MASLD and CKM (e.g., gut microbiota-liver-vascular axis, lipotoxic cardiac injury) remain incompletely elucidated, hindering optimization of holistic prevention strategies targeting multiorgan metabolic axes. Although independent associations between MASLD and CKM components (e.g., obesity, diabetes, CKD) have been established, whether MASLD exhibits unique distribution patterns and prognostic significance across the CKM spectrum remains controversial. Some studies suggest that CKM-related metabolic disturbances may overshadow MASLD’s independent risk contribution, while others posit that MASLD, via exacerbating IR, could accelerate CKM progression toward end-stage disease and adverse outcomes [15]. Furthermore, whether IR mediates the association between MASLD and CVD mortality in CKM patients remains unvalidated through causal inference models. These knowledge gaps highlight MASLD’s potential value in precision management of CKM.

This study aims to address three key questions: First, epidemiologically, to delineate the incidence and dynamic evolution of MASLD across CKM stages (1–4); second, prognostically, to validate whether MASLD elevates CVD mortality risk in CKM patients; third, mechanistically, to elucidate IR’s mediating role in the MASLD-CVD mortality association through causal mediation analysis.

## 2. Methods

### 2.1 Definitions and Data Collection

Data were derived from the National Health and Nutrition Examination Survey (NHANES) database (2009–2018), excluding CKM stage 0. Stages 1–4 of CKM were defined as the study population. NHANES is designed to examine individual-level demographic, health, and nutritional information through personal interviews and standardized physical examinations at mobile examination centers (MECs), assessing the health and nutritional status of non-institutionalized U.S. civilians[16].

To ensure accuracy and reliability, CKM staging followed the American Heart Association (AHA) framework, defined as: To ensure accuracy and reliability, CKM staging followed the AHA framework, defined as: 1) Stage 0: No CKM risk factors; 2) Stage 1: Excess or dysfunctional adiposity; 3)Stage 2: Metabolic risk factors (hypertriglyceridemia, hypertension, diabetes, metabolic syndrome) or moderate-to-high-risk CKD; 4) Stage 3: Subclinical CVD or CVD risk equivalents (high CVD risk or very high-risk CKD) within CKM syndrome; 5) Stage 4: Clinical CVD within CKM syndrome[17]. Prior CKM studies for NHANES to ensure staging accuracy (details in **Supplementary Table 1**) [18]. MASLD was defined by Fatty Liver Index (FLI) and ultrasound-based Fatty Liver Index (usFLI), with NAFLD diagnosed as usFLI ≥30 or FLI ≥ 60[19].

FLI was calculated using the formula proposed by Bedogni G et al. [20].

FLI = (e^0.953*loge (triglycerides) + 0.139*BMI + 0.718*loge (GGT) + 0.053*waist circumference - 15.745^) / (1 + e^0.953*loge (triglycerides) + 0.139*BMI + 0.718*loge (GGT) + 0.053*waist circumference - 15.745^) *100.

usFLI was calculated per Ruhl CE et al. [21].

usFLI = (e^−0.8073* non-Hispanic black + 0.3458* Mexican American + 0.0093* age + 0.6151* loge (GGT) + 0.0249* waist circumference + 1.1792*loge (insulin) + 0.8242* loge (glucose) – 14.7812^)/(1 + e^−0.8073* non-Hispanic black + 0.3458* Mexican American + 0.0093* age + 0.6151* loge (GGT) + 0.0249* waist circumference+ 1.1792*loge (insulin) + 0.8242* loge (glucose)– 14.7812^)*100

(Ethnicity variables: “Mexican American” and “non-Hispanic Black” were coded as 1 if applicable, 0 otherwise.)

Triglyceride-glucose (TyG) index was calculated as [22]: TyG = ln[triglycerides (mg/dL) × glucose (mg/dL) / 2]

Exclusion criteria: 1) Age <20 years; 2) Pregnant women; 3) Missing MASLD data; 4) Missing survival data.

NHANES protocols were approved by the National Center for Health Statistics (NCHS) Ethics Review Board, adhering to the Declaration of Helsinki. All participants provided written informed consent [23]. Definitions for smoking, alcohol use, CVD, hypertension, and diabetes are detailed in **Supplementary-table 1**.

### 2.2 Statistical Analysis

Analyses incorporated sample weights per CDC guidelines [24]. Continuous variables are expressed as mean (standard error) and categorical variables as percentages (standard error). Multiple comparisons were adjusted using the Bonferroni method. Weighted multivariate Cox regression models evaluated associations between MASLD and CVD mortality in CKM patients. Dose-response relationships between usFLI scores and CVD mortality were visualized using weighted restricted cubic splines (RCS). Sensitivity analyses included: 1) Stratified analyses by age, sex, race, hypertension (’Yes’ or ’No’), diabetes (’Yes’ or ’No’), CKD (’Yes’ or ’No’), and CVD (’Yes’ or ’No’), with interaction testing to assess subgroup heterogeneity; 2) Data analysis was performed following the exclusion of missing values to mitigate their influence; 3) Reassessment using FLI-based MASLD criteria. Mediation analysis quantified TyG’s role in the MASLD-CVD mortality relationship. Missing data were addressed using established imputation methods. All analyses were performed in R 4.3.1, with two-sided *p* <0.05 considered statistically significant.

## 3. Results

### 3.1 Baseline Characteristics

This study enrolled 49,693 patients from the NHANES database during the 2009-2018 cycle. After screening and exclusions, the study ultimately included 968 participants in CKM stage 0 and 9,093 CKM patients (encompassing stages 1, 2, 3, and 4). Among all CKM patients, the distribution was as follows: 27.9% in stage 1, 57.1% in stage 2, 4.3% in stage 3, and 10.7% in stage 4. The prevalence of MASLD among all enrolled CKM patients was 27.3%.The study population comprised 49.47% males, with a mean age of 49.89 years and an average BMI of 29.12 kg/m². The prevalence rates of hypertension, diabetes, CKD, dyslipidemia, and CVD were 40.84%, 16.72%, 15.24%, 73.59%, and 10.28%, respectively. Compared to non-MASLD individuals, CKM patients with MASLD were older, had lower eGFR, higher BMI, and a greater proportion of males (All P < 0.05). Additionally, they exhibited higher prevalence rates of hypertension, dyslipidemia, diabetes, CKD, and CVD (57.76%, 87.59%, 35.59%, 22.57%, and 16.06%, respectively) (All *P* < 0.05) (Figure 1, Table 1).

**Figure 1.**
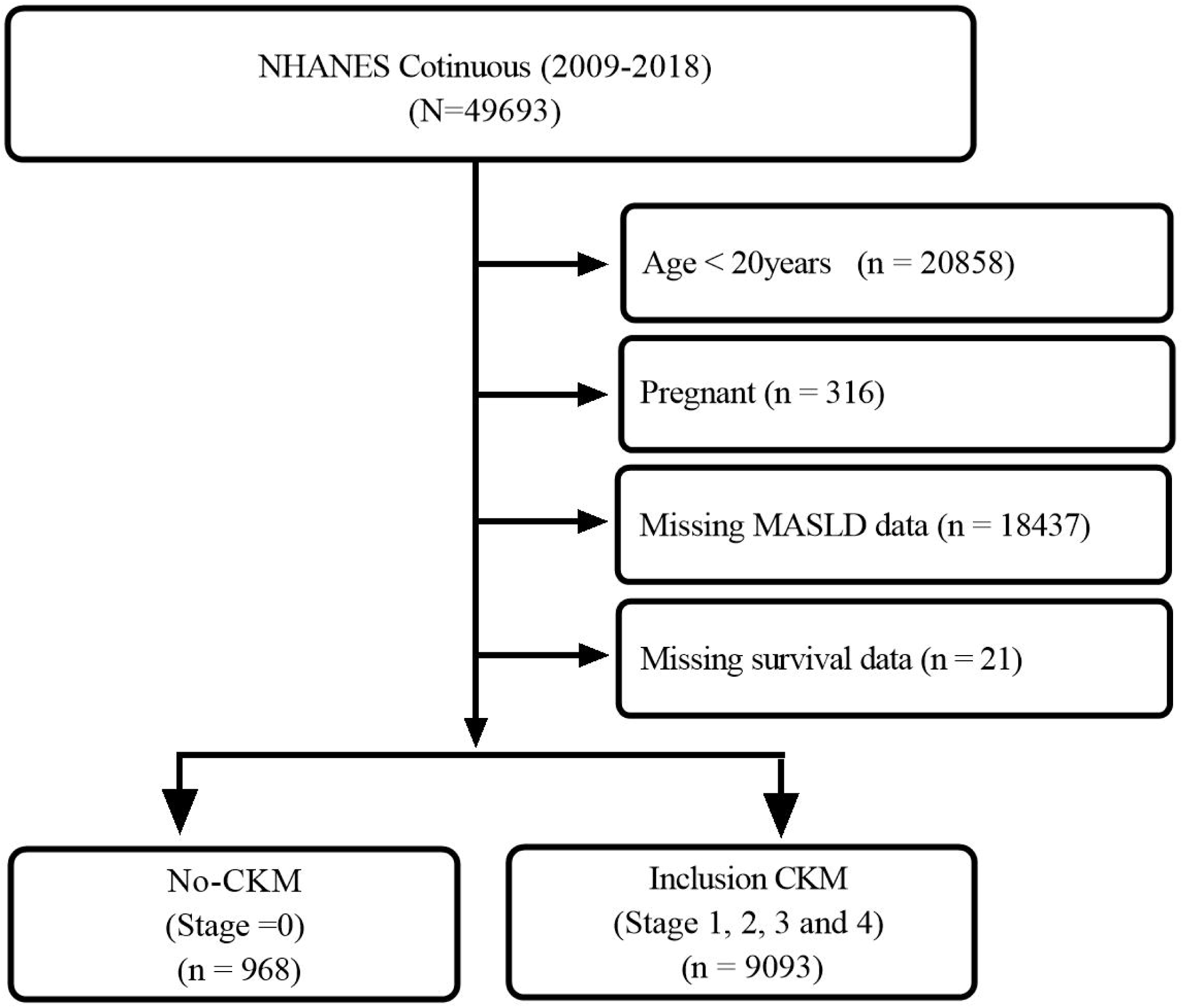
Inclusion flowchart.

**Table 1.** Baseline clinical features of enrolled individuals with CKM.

| Variable | Total<br>(N=xx) | Non-MASLD<br>(N=xx) | MASLD<br>(N=xx) | P-value |
| --- | --- | --- | --- | --- |
| Age (years) | 49.89(0.27) | 48.12(0.30) | 54.60(0.41) | < 0.001 |
| Age (years) (%) |  |  |  | < 0.001 |
| <65 | 77.56(0.02) | 80.87(0.60) | 68.73(1.33) |  |
| ≥65 | 22.44(0.01) | 19.13(0.60) | 31.27(1.33) |  |
| Sex (%) |  |  |  | < 0.001 |
| Female | 50.53(0.02) | 52.70(0.81) | 44.76(1.56) |  |
| Male | 22.44(0.01) | 19.13(0.60) | 31.27(1.33) |  |
| Ethnicity (%) |  |  |  | < 0.001 |
| Mexican American | 7.68(0.01) | 6.40(0.64) | 11.09(1.24) |  |
| Non-Hispanic Black | 11.15(0.01) | 13.12(0.99) | 5.92(0.62) |  |
| Non-Hispanic White | 66.46(0.03) | 65.27(1.56) | 69.61(1.79) |  |
| Other Hispanic | 6.02(0.01) | 6.10(0.62) | 5.82(0.67) |  |
| Other Race - Including<br>Multi-Racial | 8.68(0.01) | 9.11(0.64) | 7.56(0.69) |  |
| BMI (kg/m<sup>2</sup>) | 29.12(0.11) | 27.17(0.09) | 34.32(0.23) | < 0.001 |
| BMI (kg/m<sup>2</sup>) (%) |  |  |  | < 0.001 |
| ≤30 | 24.53(0.01) | 32.07(0.91) | 4.46(0.56) |  |
| > 30 | 75.47(0.02) | 67.93(0.91) | 95.54(0.56) |  |
| Hypertension (%) |  |  |  | < 0.001 |
| No | 59.16(0.02) | 65.52(0.92) | 42.24(1.64) |  |
| Yes | 40.84(0.02) | 34.48(0.92) | 57.76(1.64) |  |
| Alcohol user (%) |  |  |  | < 0.001 |
| No | 12.41(0.01) | 10.47(0.51) | 17.58(0.96) |  |
| Yes | 87.59(0.03) | 89.53(0.51) | 82.42(0.96) |  |
| Smoke (%) |  |  |  | 0.63 |
| No | 56.09(0.02) | 55.89(0.93) | 56.62(1.57) |  |
| Yes | 43.91(0.02) | 44.11(0.93) | 43.38(1.57) |  |
| Poverty (10,000 dollars) (%) |  |  |  | 0.01 |
| ≤1 | 19.94(0.01) | 20.19(0.94) | 19.30(1.14) |  |
| 1.1-3 | 34.86(0.01) | 33.67(1.00) | 38.04(1.40) |  |
| >3 | 45.19(0.02) | 46.15(1.42) | 42.66(1.64) |  |
| Educational level (%) |  |  |  | < 0.001 |
| College or above | 61.00(0.02) | 62.79(1.31) | 56.35(1.46) |  |
| High school or equivalent | 33.18(0.01) | 32.46(1.18) | 35.17(1.44) |  |
| Less than high school | 5.76(0.00) | 4.75(0.35) | 8.48(0.75) |  |
| Hyperlipidemia (%) |  |  |  | < 0.001 |
| No | 26.41(0.01) | 31.67(0.68) | 12.41(0.96) |  |
| Yes | 73.59(0.02) | 68.33(0.68) | 87.59(0.96) |  |
| HbA1c (%) | 5.68(0.01) | 5.51(0.01) | 6.14(0.03) | < 0.001 |
| DM (%) |  |  |  | < 0.001 |
| No | 83.28(0.03) | 90.37(0.51) | 64.41(1.25) |  |
| Yes | 16.72(0.01) | 9.63(0.51) | 35.59(1.25) |  |
| ACR (mg/g) | 35.10(2.70) | 29.21(2.69) | 50.78(6.23) | < 0.001 |
| ACR (mg/g) (%) |  |  |  | < 0.001 |
| <30 | 89.75(0.03) | 91.65(0.39) | 84.71(0.86) |  |
| (30-300) | 8.56(0.00) | 7.11(0.36) | 12.43(0.82) |  |
| ≥300 | 1.69(0.00) | 1.25(0.12) | 2.87(0.37) |  |
| eGFR (ml/min/1.73m<sup>2</sup>) | 92.95(0.36) | 94.66(0.41) | 88.41(0.55) | < 0.001 |
| e-GFR (ml/min/1.73m<sup>2</sup>) (%) |  |  |  | < 0.001 |
| ≥60 | 92.74(0.03) | 94.09(0.36) | 89.12(0.80) |  |
| <60 | 7.26(0.00) | 5.91(0.36) | 10.88(0.80) |  |
| CKD (%) |  |  |  | < 0.001 |
| No | 84.41(0.03) | 87.44(0.51) | 77.43(0.97) |  |
| Yes | 15.24(0.01) | 12.56(0.51) | 22.57(0.97) |  |
| CKM Syndrome (%) |  |  |  | < 0.001 |
| Stage1 | 27.91(0.01) | 35.32(1.00) | 8.22(0.79) |  |
| Stage2 | 57.09(0.02) | 52.80(0.92) | 68.53(1.16) |  |
| Stage3 | 4.33(0.00) | 3.46(0.22) | 6.64(0.54) |  |
| Stage4 | 10.66(0.01) | 8.42(0.53) | 16.61(1.05) |  |
| CVD (%) |  |  |  | < 0.001 |
| No | 89.72(0.03) | 91.90(0.51) | 83.94(1.01) |  |
| Yes | 10.28(0.01) | 8.10(0.51) | 16.06(1.01) |  |
| usFLI | 24.66(0.36) | 13.67(0.14) | 53.89(0.54) | < 0.001 |

CKM, Cardio-Kidney-Metabolic; MASLD, Metabolic Dysfunction-Associated Steatotic Liver Disease; BMI, Body Mass Index; ACR, albumin-to-creatinine ratio; DM, diabetes mellitus; e-GFR, estimated glomerular filtration rate; CKD, chronic kidney disease; CVD, Cardiovascular disease; usFLI, Ultrasound-based Fatty Liver Index.

### 3.2 Incidence of MASLD in CKM

Among the included CKM patients, an epidemiological analysis of MASLD was conducted. In those with coexisting MASLD, the distribution across CKM stages 1, 2, 3, and 4 was 8.22%, 68.53%, 6.64%, and 16.61%, respectively **(Table 1)**. Across the five survey cycles (2009–2010, 2011–2012, 2013–2014, 2015–2016, and 2017–2018), the prevalence of MASLD was 28.51%, 26.99%, 24.62%, 28.33%, and 28.09%, respectively. No significant temporal trend in MASLD incidence was observed (*p* > 0.05) **(Table 2)**.

**Table 2.** Magnitude of Changes in Mean MASLD for Subsequent National Health and Nutrition Examination Survey Cycles in individual with CKM.

| Survey years | MASLD | Difference | P value for difference in BRI<sup>b</sup> | Overall P value for trend<sup>c</sup> |
| --- | --- | --- | --- | --- |
|  | Mean (SE) [95% CI] |  |  |  |
| 2009-2010 | 28.51(1.34) | 0[Reference] | NA | 0.38 |
| 2011-2012 | 26.99(1.76) | -1.52 | 0.5 |  |
| 2013-2014 | 24.62(1.58) | -3.89 | 0.07 |  |
| 2015-2016 | 28.33(1.49) | -0.18 | 0.93 |  |
| 2017-2018 | 28.09(1.62) | -0.42 | 0.84 |  |

Abbreviations: MASLD, Metabolic Dysfunction-Associated Steatotic Liver Disease; NA, not applicable.
<sup>a</sup> Nationally representative estimates of the nonpregnant US population aged 20 years or more. Estimates were nationally representative through the use of survey weights.
<sup>b</sup> P value for difference in mean BRI was calculated using the linear combinations of parameters.
<sup>c</sup> P value for overall trend was calculated using the Mann-Kendall trend test.

The prevalence of MASLD among individuals with CKM stages 1, 2, 3, and 4 was 8.04%, 32.78%, 41.90%, and 42.55%, respectively. The prevalence of MASLD increased progressively from CKM stage 1 to stage 4 (*P* < 0.001). Furthermore, compared to CKM stage 1 individuals, those in CKM stages 2, 3, and 4 exhibited significantly higher MASLD prevalence rates (All *P* < 0.001) **(Table 3)**. In addition, among CKM patients with different age (’<65’ years, ’≥65’ years), gender (’Female’, ’Male’), and race (’Mexican American’, ’Non-Hispanic Black’, ’Non-Hispanic White’, ’Other Hispanic’, ’Other Race - Including Multi-Racial’), the prevalence of MASLD showed a gradually increasing trend with the progression of CKM (*P* < 0.001). Moreover, compared with individuals in CKM stage 1, the prevalence of MASLD was higher in individuals in CKM stages 2, 3, and 4 (All *P* < 0.001) **(Table 3)**.

**Table 3.**
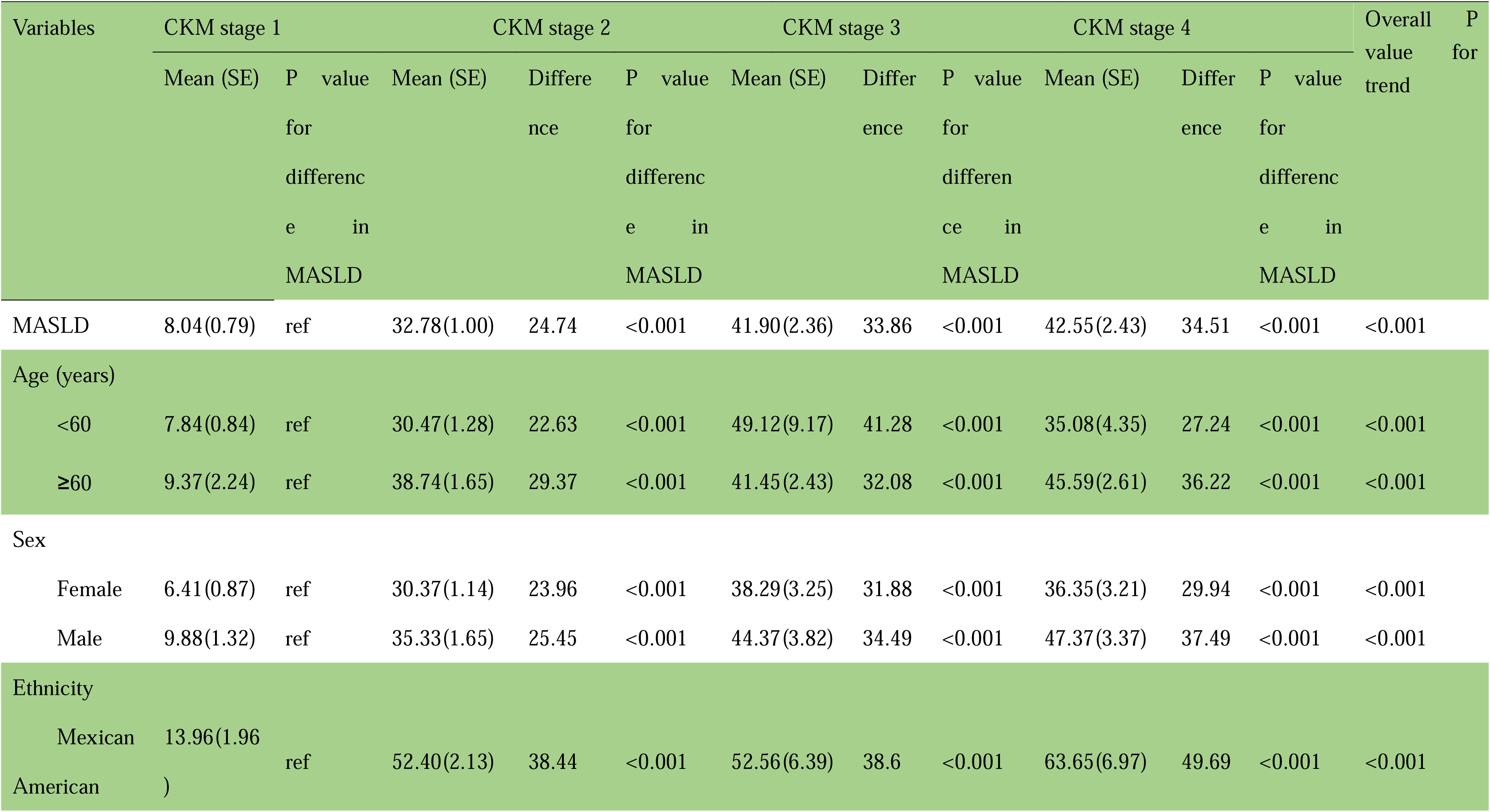
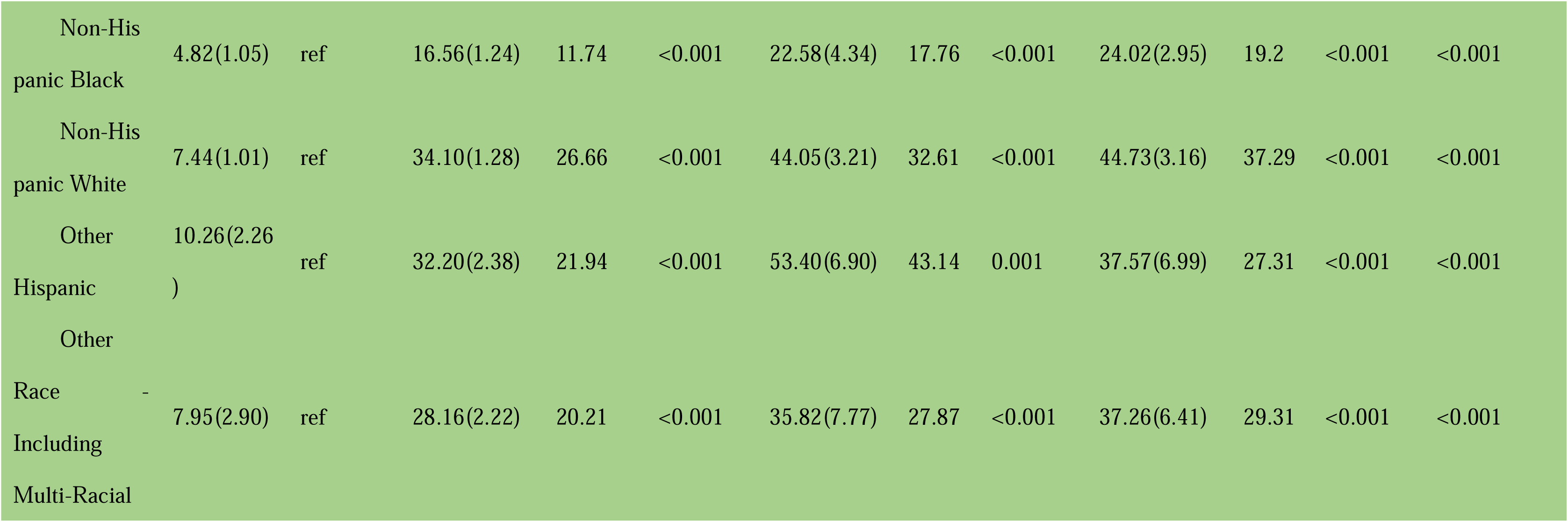
Cox-regression analysis of risk factors for CVD mortality in individuals with CKM.

### 3.3 Association Between MASLD and CVD Mortality Risk in CKM Patients

#### 3.3.1 Exploration of Nonlinear Relationships

We employed RCS to model the dose-response relationship between the usFLI score and CVD mortality risk in CKM patients. Overall, CVD mortality risk increased linearly with higher usFLI scores (nknots = 3, *P* for overall < 0.001, Non-line *P* = 0.38) **(Figure 2 A)**. When stratified by sex, a significant linear trend of increasing CVD mortality risk with elevated usFLI scores was observed in the male subgroup (nknots = 3, *P* for overall = 0.001,Non-line *P* = 0.71): In the female subgroup, no significant dose-response relationship was observed (nknots = 3, *P* for overall = 0.33, Non-line *P* = 0.55) **(Figure 2 B)**. Age-stratified analyses revealed a significant linear increase in CVD mortality with higher usFLI scores across all age groups: ≤ 65 years group(nknots = 3, *P* for overall < 0.001, Non-line *P* = 0.77), > 65 years group (nknots = 3, *P* for overall = 0.004,Non-line *P* = 0.24): However, the ≤65-year subgroup exhibited a steeper slope **(Figure 2 C)**. When stratified by hypertension status (’Yes’ or ’No’), CKM patients with hypertension exhibited a more pronounced linear increase in CVD mortality risk with rising usFLI scores **(Figure 2 D)**. In diabetes-stratified analyses, a U-shaped relationship emerged between usFLI scores and CVD mortality among CKM patients with DM (nknots = 3, *P* for overall < 0.001, Non-line *P* = 0.04); specifically, CVD mortality risk increased with usFLI scores >30. In contrast, no dose-response relationship was observed in non-DM patients **(Figure 2 E)**. Stratified by CKD (’Yes’ or ’No’) and CVD (’Yes’ or ’No’) status: Individuals without CKD (nknots = 3, *P* for overall < 0.001, Non-line *P* = 0.21) or CVD (nknots = 3, *P* for overall < 0.001, Non-line *P* = 0.45) exhibited a significant linear association between usFLI scores and CVD mortality. In contrast, no dose-response relationship was observed in individuals with pre-existing CKD or CVD **(Figure 2 F, G)**. Stratified by body mass index (BMI), a significant linear association was observed between usFLI scores and CVD mortality among individuals with BMI > 30 kg/m²(nknots = 3, *P* for overall = 0.003, Non-line *P* = 0.98) **(Figure 2 H)**. In analyses stratified by CKM stages, a significant linear association was observed between usFLI scores and CVD mortality specifically in CKM stage 2 patients (nknots = 3, *P* for overall = 0.01, Non-line *P* = 0.77). No dose-response relationship was detected in other CKM stages (All *P* for overall > 0.05) **(Figure 2 I)**.

**Figure 2.**
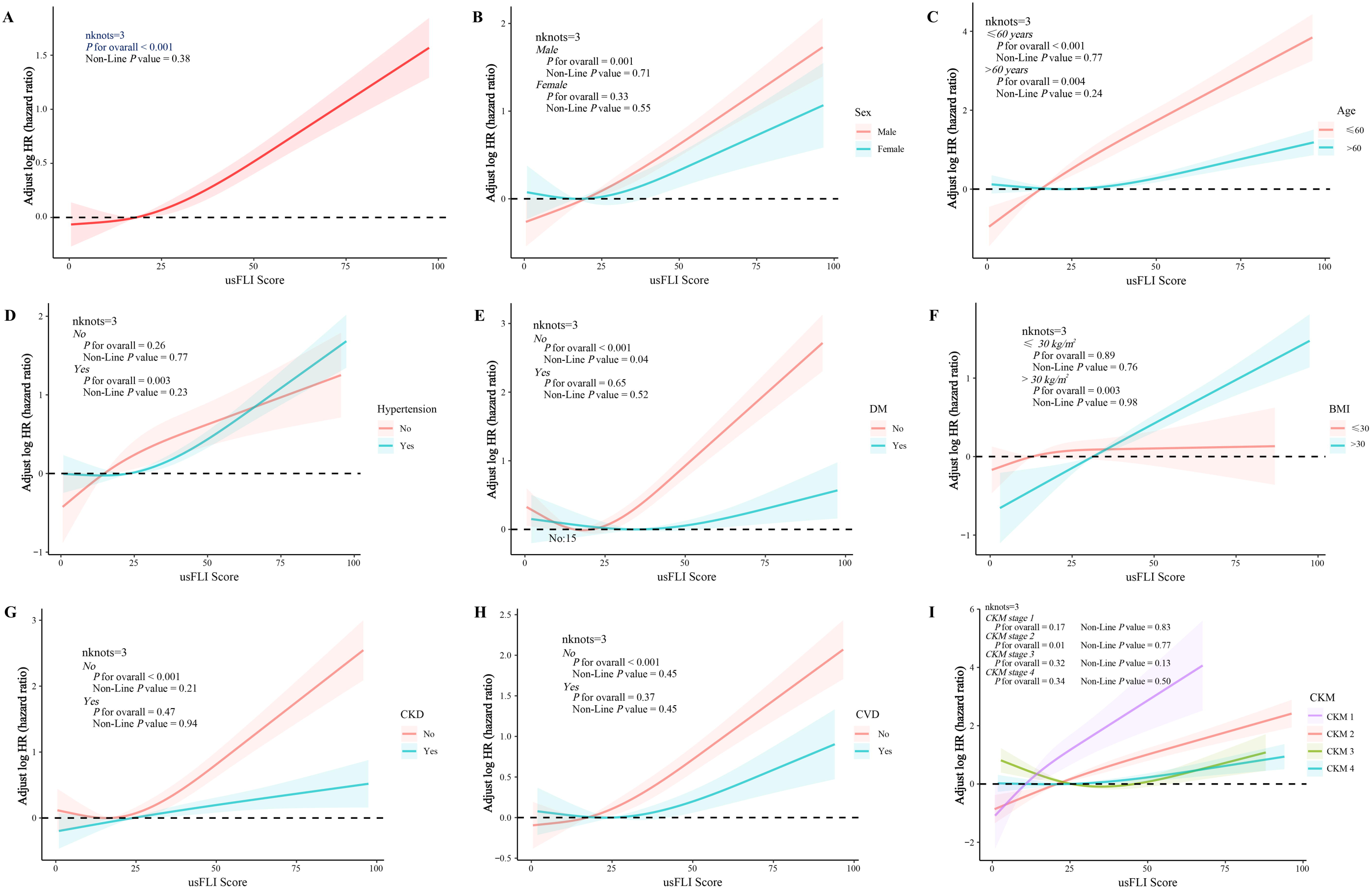
The relationship between MASLD and the prognosis of individuals with CKM through Kaplan-Meier survival analysis. MASLD, Metabolic Dysfunction-Associated Steatotic Liver Disease; CKM, Cardiovascular-kidney-Metabolic.

#### 3.3.2 Multivariate Cox Regression Analysis

The relationship between MASLD and the risk of CVD mortality in CKM patients was evaluated through a multivariate Cox regression model. After adjusting for age (‘<65’ years, ‘≥65’ years), gender (‘Female’, ‘Male’), race (‘Mexican American’, ‘Non-Hispanic Black’, ‘Non-Hispanic White’, ‘Other Hispanic’, ‘Other Race - Including Multi-Racial’), BMI (<30kg/m^2^, ≥30 kg/m^2^), smoke (’Yes’ or ’No’), alcohol use (’Yes’ or ’No’), education (‘College or above’, ‘High school or equivalent’, ‘Less than high school’), poverty-income ratio (PIR) (‘$0-10k’, ‘$11-30k’, ‘,$30k’), and CVD (’Yes’ or ’No’), the results demonstrated that CKM patients with MASLD had a 63% higher risk of CVD mortality compared to those without MASLD (HR = 1.63; 95% CI, 1.05 - 2.52, *P* = 0.03) **(Figure 3)**.

**Figure 3.**
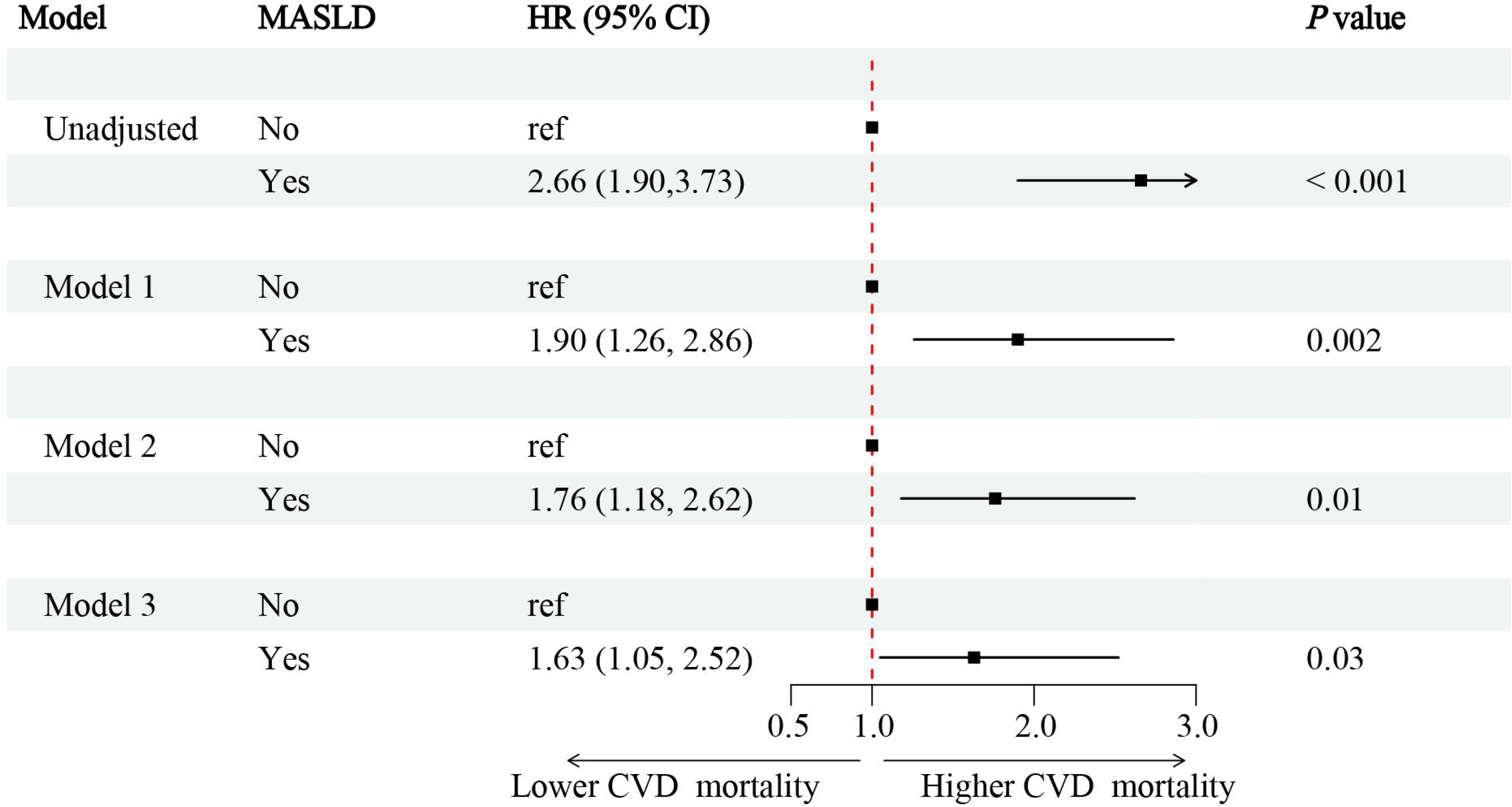
Association between MASLD and CVD mortality in individuals with CKM. **Model 1** adjusted for baseline age (‘<65’ years, ‘≥65’ years), gender (‘Female’, ‘Male’), ethnicity (‘Mexican American’, ‘Non-Hispanic Black’, ‘Non-Hispanic White’, ‘Other Hispanic’, ‘Other Race - Including Multi-Racial’), BMI (<30kg/m^2^, ≥30 kg/m^2^); **Model 2** adjusted for covariates in model 1 plus smoke (‘Yes’ or ‘No’), alcohol use (‘Yes’ or ‘No’), education (‘College or above’, ‘High school or equivalent’, ‘Less than high school’), poverty (‘0-1’, ‘1.1-3’, ‘,3’); **Model 3** adjusted for covariates in model 2 plus CVD (‘Yes’ or ‘No’). MASLD, Metabolic Dysfunction-Associated Steatotic Liver Disease; CKM, Cardiovascular-kidney-Metabolic; HR, Hazard ratio; CI, Confidence interval; BMI, Body Mass Index; CVD, Cardiovascular disease.

### 3.4 Mediation Analysis

MASLD demonstrated a significant overall effect on CVD mortality in CKM patients, with a total effect (TE) of 0.013(TE = 0.013; 95% CI, 0.006 - 0.021, *P* < 0.001). In terms of mediation effects, MASLD may influence CVD mortality in CKM patients by affecting the mediator TyG, with an average causal mediation effect (ACME) of TyG being 0.005 (ACME = 0.005; 95% CI, 0.002 - 0.009, *P* < 0.001),The mediation effect accounted for 40.5% of the total effect, indicating that 40.5% of MASLD’s total effect on CVD mortality in CKM patients was mediated by TyG. Meanwhile, the direct effect of MASLD on CVD mortality in CKM patients (the portion not mediated by TyG) had an ACME of 0.008 (ACME = 0.008; 95% CI, 0.000 - 0.015, *P* = 0.04),This indicates that even after accounting for the mediating role of TyG, MASLD still exerts a significant direct effect on CVD mortality (**Supplementary -table 2,3**; **Figure 4**).

**Figure 4.**
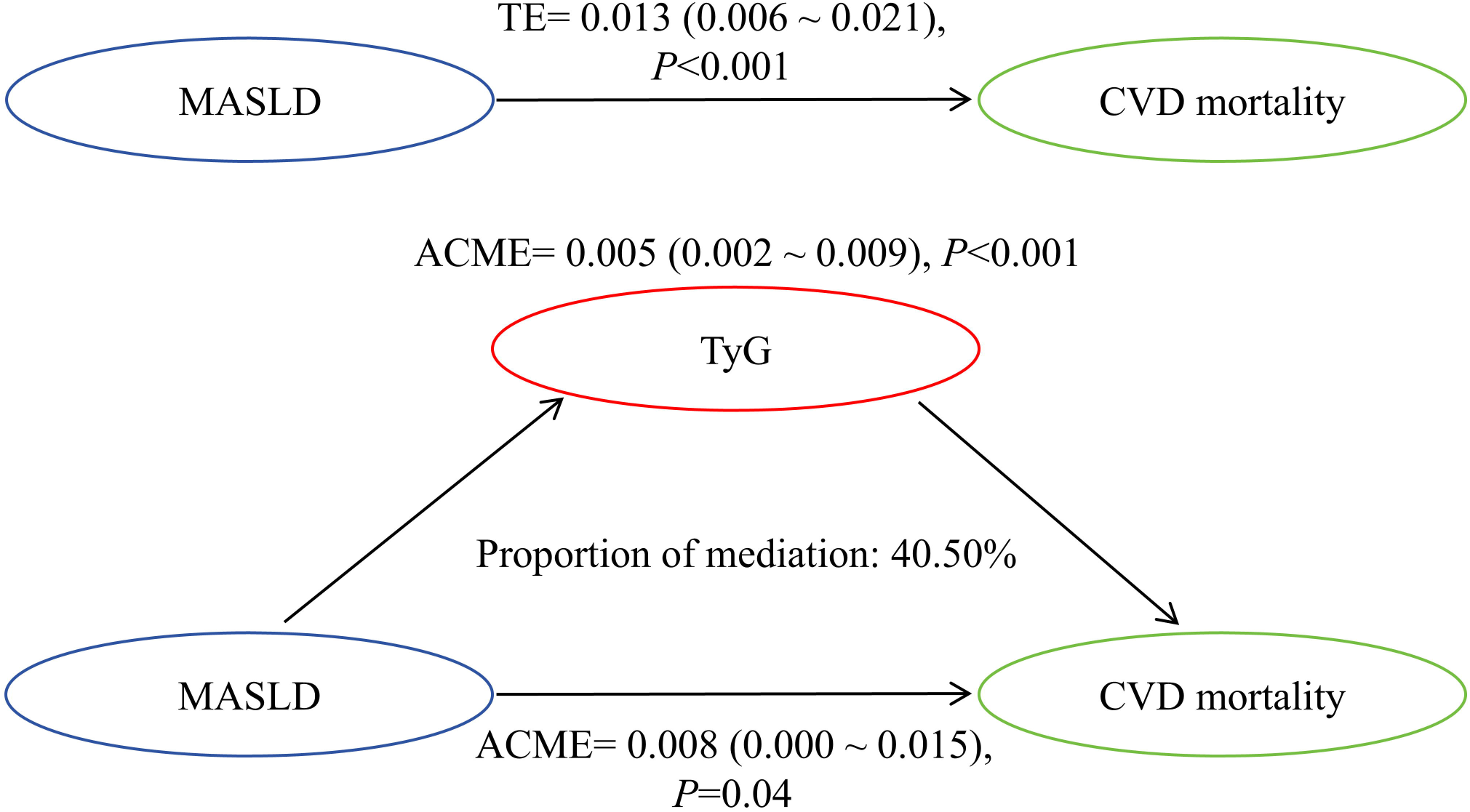
Mediation effect of TyG index between MASLD and CVD mortality. Triglyceride-glucose index; MASLD, Metabolic Dysfunction-Associated Steatotic Liver Disease; CVD, Cardiovascular disease.

### 3.5 Sensitivity Analysis

#### 3.5.1 Stratified Analysis

Stratified analysis by age (‘<65’ years, ‘≥65’ years), Sex (‘Female’, ‘Male’), BMI (<30kg/m^2^, ≥30 kg/m^2^), Hypertension (’Yes’ or ’No’), DM (’Yes’ or ’No’), CKD (’Yes’ or ’No’), CVD (’Yes’ or ’No’) revealed significant interaction effects between MASLD and age, DM, CKD, CVD, as well as CKM stages (All *P* for interaction < 0.05). In the <65-year-old age group, the presence of MASLD increased the risk of CVD mortality by 6.39-fold in CKM patients (HR = 7.39; 95% CI, 3.21 - 17.01, *P*< 0.001): In patients with MASLD but without comorbid DM, CKD, or CVD, the risk of CVD mortality was significantly elevated, showing increases of 1.17-fold (HR = 2.17; 95% CI, 1.12–4.17; *P* = 0.02), 1.92-fold (HR = 2.92; 95% CI, 1.38–6.19; *P* = 0.01), and 1.46-fold (HR = 2.46; 95% CI, 1.38–4.41; *P* = 0.002), respectively. Among patients at different CKM stages, those at CKM stage 1 with MASLD exhibited a 5.51-fold increased risk of CVD mortality (HR = 6.51; 95% CI, 1.19-35.68; *P* = 0.03), while those at CKM stage 2 with MASLD showed a 3.65-fold increased risk (HR = 4.65; 95% CI, 1.92-11.26; *P* < 0.001).MASLD showed no significant interaction effects with gender (’Female’ or ’Male’), BMI (<30 kg/m ² or ≥ 30 kg/m ²), hypertension (’Yes’ or ’No’), or other factors in relation to CVD risk (All *P* for interaction > 0.05) (Supplementary-table 4,Figure 5) .

**Figure 5.**
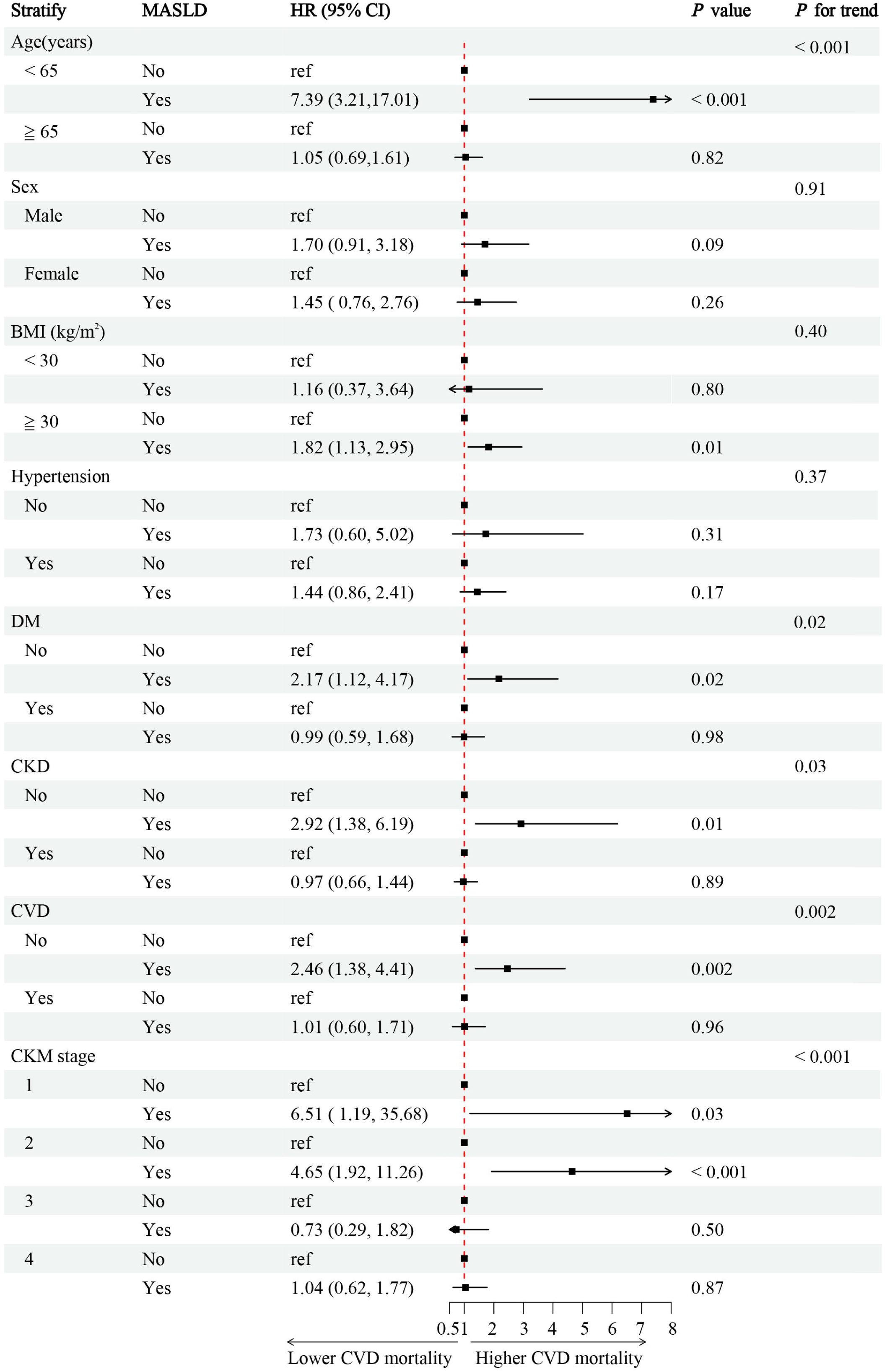
Stratified analysis. adjusted for baseline age (‘<65’ years, ‘≥65’ years), gender (‘Female’, ‘Male’), ethnicity (‘Mexican American’, ‘Non-Hispanic Black’, ‘Non-Hispanic White’, ‘Other Hispanic’, ‘Other Race - Including Multi-Racial’), BMI (<30kg/m^2^, ≥30 kg/m^2^), smoke (‘No’ or ‘Yes’), alcohol use (‘No’ or ‘Yes’), education (‘College or above’, ‘High school or equivalent’, ‘Less than high school’), poverty (‘0-1’, ‘1.1-3’, ‘,3’). HR, Hazard ratio; CI, Confidence interval; BMI, Body Mass Index; CKD, chronic kidney disease; DM, diabetes mellitus; CVD, Cardiovascular disease; CKM, Cardiovascular-kidney-Metabolic.

#### 3.5.2 Missing Data Analysis Using Complete Case Analysis

After performing censoring processing for missing data, we re-evaluated the association between MASLD and CVD mortality risk in CKM patients to ensure result stability and validate hypothesis rationality, thereby enhancing conclusion reliability. Following multivariate adjustment, Cox regression analysis demonstrated that using non-MASLD CKM patients as reference, the MASLD group exhibited 1.84-fold higher CVD mortality risk compared to the non-MASLD group (HR = 1.84; 95% CI, 1.14 - 2.96, *P* = 0.012) **(Supplementary-table 5)**.

#### 3.5.3 Evaluation of the relationship between MASLD and the risk of CVD mortality using the FTI as the diagnostic criterion for MASLD

After applying alternative diagnostic criteria for MASLD, we re-evaluated its association with CVD mortality risk in CKM patients. Following multivariable adjustment, Cox regression analysis revealed that compared to the non-MASLD group, CKM patients with MASLD exhibited a 68% increased risk of CVD mortality (HR = 1.68; 95% CI, 1.07 - 2.63, *P* = 0.025) **(Supplementary-table 6)**.

## 4. Discussion

This study utilized NHANES data to elucidate the epidemiological characteristics of MASLD in CKM patients, demonstrating a progressive increase in MASLD risk with advancing CKM stages. Furthermore, our findings revealed a significant association between MASLD and elevated CVD mortality risk in CKM patients. Through causal mediation analysis, we identified that insulin resistance (TyG index) mediated 40.5% of the MASLD-associated CVD mortality risk. These results provide direct population-based evidence supporting the “cardio-hepato-kidney-metabolic axis” hypothesis and its underlying pathological mechanisms.

Kalligero et al. analyzed that the prevalence rate of MASLD among American adults was 32.45%[25]. In the CKM patients included in this study, the incidence of MASLD was 27.3%, which is lower than the prevalence rate in the general population. This may be because some CKM patients died of CVD or kidney diseases before progressing to the clinical diagnosis stage of MASLD [26]. Zhang et al. [27] conducted a statistical study on the global burden of metabolic diseases from 1990 to 2021 and found that there was no significant change in MASLD over time. Consistent with our findings, the analysis of five NHANES cycles revealed relatively stable MASLD prevalence rates over time, suggesting its persistently high disease burden that warrants continued research for incidence reduction. Notably, compared to non-MASLD individuals, CKM patients with MASLD exhibited distinct clinical characteristics including: (1) advanced age, (2) lower eGFR levels, (3) higher BMI values, and (4) greater male predominance - all aligning with established epidemiological patterns reported in multiple cohort studies [28, 29]. Moreover, we identified a significant positive correlation between MASLD prevalence and advancing CKM stages, with rates escalating from 8.04% in stage 1 to 42.55% in stage 4 patients. This progressive pattern suggests that CKM syndrome progression is closely associated with exacerbated hepatic steatosis and metabolic dysfunction. The underlying mechanism may involve the diagnostic prerequisite for MASLD requiring ≥ 3 of 5 metabolic syndrome (MetS) components (including increased waist circumference, elevated blood pressure, raised fasting triglycerides/glucose, or reduced HDL-cholesterol), where each additional MetS component elevates CVD risk, CVD mortality, and all-cause mortality by approximately 2.4-fold [30]. Consequently, higher CKM stages, reflecting greater MetS component accumulation, correspondingly increase MASLD susceptibility.

Patients with MASLD demonstrate significantly higher comorbidity rates of hypertension, diabetes, CKD, and CVD. Thus, MASLD is not merely a metabolic “byproduct” but may also serve as a systemic “driver” for multi-organ damage. Hepatic fat accumulation exacerbates systemic endothelial dysfunction through free fatty acid (FFA) release and pro-inflammatory cytokine production, thereby accelerating atherogenesis. A large-scale study (n=234,488) revealed that significant liver fibrosis correlates with increased CKD prevalence, while MASLD phenotypes featuring steatotic liver disease (SLD) plus any two MetS components confer the highest CKD risk(OR 1.53,95% CI: 1.35–1.74, P < 0.0001) [31]. Current evidence indicates that MASLD patients face a 2- to 5-fold increased risk of developing T2DM, along with a 40%–60% elevated risk of CVD events (e.g., myocardial infarction, stroke) compared to non-MASLD populations [32, 33]. Current evidence indicates that MASLD patients face a 2- to 5-fold increased risk of developing T2DM, along with a 40%–60% elevated risk of CVD events (e.g., myocardial infarction, stroke) compared to non-MASLD populations.

Multiple studies demonstrate a significant association between MASLD and increased CVD mortality in the general population. A landmark NAFLD cohort study revealed a linear escalation in CVD mortality risk with elevated FLI scores (HR, 95% CI: FLI 30-59 = 1.18, 1.16-1.20; FLI ≥ 60 = 1.61, 1.56-1.65)[34]. A separate prospective cohort study of 15,784 participants from the EPIC (European Prospective Investigation into Cancer and Nutrition) study demonstrated robust positive associations between MASLD and all-cause, cancer-related, and CVD mortality. Notably, the association with CVD mortality exhibited a significant dose-response relationship with increasing FLI scores [35].This study demonstrates that within the CKM population, patients developing MASLD face significantly elevated CVD mortality risk, with a pronounced dose-dependent increase corresponding to MASLD severity (usFLI scores). After multivariable adjustment, MASLD was associated with a 63% higher CVD mortality risk. Sensitivity analyses employing alternative diagnostic criteria (FTI) and censoring approaches consistently maintained statistically significant hazard ratios, confirming the robustness of these findings. The elevated CVD mortality risk in CKM patients with MASLD likely results from the interplay of metabolic inflammation, multi-organ crosstalk, and suboptimal clinical management [36]. These findings underscore the imperative for heightened clinical vigilance toward MASLD in the CKM population, necessitating both early precision screening and timely risk factor modification.

The results of the RCS analysis and stratification analysis showed that in the early stage of CKM and in patients without comorbid CVD, DM, or CKD, the correlation between MASLD and CVD mortality was stronger, and the risk effect of the usFLI score was more significant. This indicates that MASLD may have a stronger predictive role for CVD events in these patients. The increase in risk was more significant in the group of patients aged ≤65 years. This may be because the comorbid diseases in the population aged > 65 years can overshadow the contribution of MASLD. However, the mechanism underlying the higher risk of CVD mortality in the younger subgroup remains unclear.

Similarly, the association between MASLD and CVD mortality was more significant in patients with CKM stages 1-2. In contrast, the risk of CVD mortality in patients with CKM stages 3-4 may be mainly related to mortalitys caused by diabetes and related complications of CKD, and the contribution of MASLD may be reduced due to “competitive risks”. These results are mainly due to the fact that risk factors such as an increase in comorbidities and deterioration of renal function may mask the independent contribution of MASLD. Therefore, especially in the early stage of CKM, it is necessary to include the observation of MASLD. MASLD should be intervened as early as possible in CKM patients, and more attention should be paid to the concept of cardio-renal-hepatic metabolic syndrome. This will help reduce the risk of CVD mortality.

Recent studies have revealed a two-way vicious cycle mechanism between IR and MASLD. IR promotes the ectopic deposition of liver fat and lipotoxicity, which induces damage to liver cells, and thus drives the occurrence and development of MASLD [37]. In patients with MASLD, the accumulation of liver lipids will further inhibit the tyrosine phosphorylation of IR substrate through the release of mediators such as free fatty acids, pro-inflammatory cytokines, and exosomal miRNAs, leading to disorders of insulin signaling in peripheral tissues [38]. It is worth noting that this IR-MASLD cycle is not limited to the liver, and it may also amplify the systemic risk of CVD through metabolites and inflammatory mediators.

Insulin resistance is an important factor contributing to CVD mortality in patients with metabolic diseases. Recent studies have shown that the liver-derived IR associated with MASLD can mediate CVD mortality by increasing the TyG index. A cohort study involving 2,018 patients with cardiometabolic syndrome (CMS) found that for every 1-unit increase in the TyG index, the risk of CVD mortality increased by 39%. The literature indicates that insulin resistance increases the risk of mortality in patient s[39]. In patients with CKM, the TyG index has been proven to be an independent predictor of CVD events. A study by Chen et al. involving 10,390 participants found that a high level of the TyG index was significantly associated with the CVD mortality rate in patients with MASLD [40]. Through causal mediation analysis, we found that the TyG index has a mediating effect between MASLD and CVD mortality, and it can explain more than 40.5% of the strength of the association. This suggests that in the case of CKM patients with MASLD, hepatic steatosis further amplifies the systemic metabolic disorder by exacerbating liver-derived insulin resistance, thus additionally increasing the risk of CVD mortality. The mechanism underlying the mediating effect of this strong association has not been fully elucidated. The multi-organ metabolic disorder driven by liver-derived insulin resistance may be the key mechanism for the increased CVD mortality rate. In patients with MASLD, the accumulation of liver fat inhibits the insulin signaling pathway through lipotoxic metabolites and pro-inflammatory factors (such as IL-6 and TNF-α), leading to an increase in hepatic glucose output and impairment of peripheral glucose uptake [41].

At the same time, it is accompanied by an increase in the secretion of very low-density lipoprotein (VLDL) and a decrease in the activity of lipoprotein lipase. These factors jointly lead to an increase in fasting blood glucose and triglyceride levels, thus significantly raising the TyG index [42]. Recent studies have further revealed that hepatocyte-derived exosomal miR-34a may regulate the above-mentioned metabolic pathway by targeting the PPARα gene. The elevated TyG index, through IR, induces a decrease in the synthesis of nitric oxide in vascular endothelium and an increase in the production of reactive oxygen species [43], and promotes endothelial dysfunction and atherosclerosis. At the same time, the increase in small and dense low-density lipoproteins (sdLDL) and the impairment of high-density lipoprotein (HDL) function in an IR environment further accelerate plaque formation [44, 45]. Therefore, targeted intervention strategies for IR may significantly reduce the risk of CVD.

In addition, synthesizing the above research findings, this study provides new insights for the clinical management of CKM syndrome. We recommend incorporating MASLD into the CKM staging system to optimize risk stratification. For patients with CKM stages 1-2, routine monitoring of hepatic steatosis using liver ultrasound or FIB-4 scores should be implemented to early identify reversible metabolic abnormalities. In patients with CKM stages 3-4, although the independent predictive value of MASLD diminishes, its synergistic effects with diabetes and CKD require individualized therapeutic interventions. For high-risk subgroups (males, ≤65 years old, BMI >30), prioritized intensive screening and stratified management should be adopted, along with community education programs. Regarding pharmacotherapy, agents with dual metabolic-modulating and anti-inflammatory effects (e.g., GLP-1 receptor agonists, SGLT2 inhibitors) should be prioritized, and the potential benefits of liver-targeted therapies (e.g., FXR agonists) on cardiorenal outcomes should be explored. Furthermore, a multidisciplinary collaborative team should be established, including specialists from cardiology, endocrinology, hepatology, nephrology, nutrition, and nursing, to regularly assess liver enzymes, cardiac ultrasound, and renal function, thereby achieving integrated “cardio-hepato-kidney-metabolic” management.

However, the limitations of the study also need to be carefully considered. Firstly, the diagnosis of MASLD relies on indirect indicators such as the FLI and the usFLI. Their sensitivity is lower than that of liver biopsy or Magnetic Resonance Imaging-Proton Density Fat Fraction (MRI-PDFF), which may underestimate the impact of liver fibrosis or inflammatory activity. Secondly, although the main confounding factors have been adjusted for, unmeasured variables (such as gut microbiota, genetic susceptibility, etc.) may still have residual confounding effects, and the limited representativeness of the population (only including the American population) also restricts the comprehensiveness of the conclusions. Finally, as a surrogate marker, the TyG index cannot fully characterize the specific pathways of the “cardio-hepato-reno-metabolic” axis, and key molecular mechanisms need to be elucidated through metabolomics or animal experiments.

## 5. Conclusion

This study confirms that MASLD is highly prevalent in patients with CKM syndrome. Its prevalence increases with the progression of CKM, and it independently increases the risk of CVD mortality. Stratified analysis suggests that in the early stage of CKM, the relationship between MASLD and the risk of CVD mortality in CKM patients may be more significant. Among them, the exacerbation of IR caused by MASLD plays a crucial mediating role in its association with CVD mortality. Integrating the liver phenotype into the CKM staging system and targeting the liver-derived IR pathway may become a key strategy to break the vicious cycle of cardio-hepato-kidney metabolism. Future research can track the dynamic interaction mechanism between MASLD and CKM staging through multicenter longitudinal cohorts, construct an early warning system by combining various novel markers, and further establish a whole-course management system for CKM based on individualized risks.

## Data Availability

All data produced in the present study are available upon reasonable request to the authors

## Funding

The Science and technology fund of Chengdu Medical College (CYZYB22-02).

The research fund of Sichuan Medical and Health Care Promotion institute (KY2022QN0309).

Sichuan Provincial Medical Association Youth Innovation Project (Q23021).

Health Commission of Sichuan Province Medical Science and Technology Program (24QNMP100).

Sichuan Provincial Geriatric Clinical Medicine Research Center (24LHLNYX1-48).

## Author Contributions

Conception and design of the study: Zhuoxing Li, Qianyu Yang, Mao Xiao, Xue Zhang, Xiunan Liu, Yanyi Deng, Yun Sun, Xiang Xiao;

Acquisition and analysis of data: Zhuoxing Li, Qianyu Yang, Yun Sun, Xiang Xiao;

Drafting the manuscript or figures: Zhuoxing Li, Mao Xiao, Xue Zhang, Yanyi Deng, Yun Sun, Xiang Xiao.

## Data Availability

Some or all datasets generated or analyzed during the current study are not publicly available. However, they can be obtained from the corresponding author upon reasonable request.

## Acknowledgments

The authors express their gratitude to all participants of this study for their significant contributions.

## Ethical Approval

The study protocol adhered to the ethical standards set forth in the 1964 Declaration of Helsinki and its subsequent amendments. Approval was obtained from the National Committee for Ethical Review of Health Statistics Research, and all participants provided signed informed consent.

## Declarations

The authors state that there are no known financial conflicts of interest or personal relationships that could have influenced the research presented in this paper.

