## Supplementary table 1-6 for "Stratified Burden of MASLD in Cardiovascular-Kidney-Metabolic syndrome: Stage-Dependent Prevalence and Insulin Resistance-Driven Cardiovascular Mortality"

**Supplementary table 1. Definitions/criteria of some diagnoses**

| Variables | Definitions/criteria |
| --- | --- |
| Smoker | Smoking more than 100 cigarettes in previous and now. |
| Alcohol user ^[1]^ | ≥2 drinks per day for females, ≥3 drinks per day for males, or binge drinking ≥2 days per month.  Binge drinking (≥4 drinks on the same occasion for females, ≥5 drinks on the same occasion for males) on 5 or more days per month. |
| Hypertension ^[2]^ | (1) Self-reported hypertension diagnosis, (2) Use of anti-hypertensive medication, (3) Average systolic blood pressure (SBP) > 140 mmHg, (4) Average diastolic blood pressure (DBP) > 90 mmHg, meet any of the above conditions. |
| Hyperlipidemia | (1) Triglyceridemia ≥ 150 mg/dl; (2) Hypercholesterolemia: a) total cholesterol ≥ 200 mg/dl, b) low-density lipoprotein ≥ 130 mg/dl), c). high-density lipoprotein (< 40 mg/dl, male; < 50 mg/dl, female), meet any of the above conditions; (3) Use of lipid-lowering drugs; meet any of the above conditions. |
| Chronic Kidney Disease (CKD) | Individuals with an albumin-to-creatinine ratio (ACR) higher than 30 mg/g and/or an estimated glomerular filtration rate (eGFR) lower than 60 mL/min/1.73 m^2^ were defined as CKD patients ^[4]^. |
| Cardiovascular Disease | Coronary heart disease; congestive heart failure; heart attack; stroke; angina. |
| Metabolic Dysfunction-Associated Steatotic Liver Disease (MASLD) ^[5]^ | At least 1 out of 5:   1. BMI≥25 kg/m2 [23 Asia] OR WC>94 cm (M) 80 cm (F) OR ethnicity adjusted equivalent； 2. Fasting serum glucose≥5.6 mmol/L[100 mg/dL] OR 2-hour post-load glucose levels≥7.8 mmol/L [≥140 mg/dL] OR HbA1c≥5.7% [39 mmol/L] OR type 2 diabetes OR treatment for type 2 diabetes； 3. Blood pressure ≥130/85 mmHg OR specific antihypertensive drug treatment； 4. Plasma triglycerides ≥1.70 mmol/L [150 mg/dL] OR lipid lowering treatment； 5. Plasma HDL-cholesterol ≤ 1.0 mmol/L [40 mg/dL] (M) and s 1.3 mmol/L [50 mg/dL] (F) OR lipid lowering treatment. |
| Cardio-Kidney-Metabolic (CKM) Syndrome ^[6]^ | We define CKM stage 0 as non-CKM, and CKM stages 1, 2, 3, and 4 as CKM.  **Stage 0: No CKM health risk factors**  CKM Stage 0 included participants with normal body mass index (BMI) (<23 kg/m2 for individuals with Asian ethnicity and <25 kg/m2 for other racial and ethnic groups), normal waist circumference (<80 and <90 cm for women and men with Asian race, respectively, and <88 and <102 cm for women and men in all other race and ethnicity categories, respectively) who did not meet criteria for the other stages;  **Stage 1: Excess and/or dysfunctional adiposity**   - CKM Stage 1 identified individuals with an elevated BMI (≥23 kg/m2 for individuals with Asian race and >25 kg/m2 for all other race and ethnic groups), an elevated waist circumference (≥80 and ≥90 cm for women and men with Asian race, respectively, and ≥88 and ≥102 cm for women and men in other race and ethnicity categories, respectively), or prediabetes (defined as a glycated hemoglobin of 5.7% to <6.5% or a fasting blood glucose of 100 mg/dL to <126 mg/dL);   **Stage 2: Metabolic risk factors and CKD**  CKM Stage 2 identified participants with metabolic risk factors or moderate-to-high-risk CKD per Kidney Disease Improving Global Outcomes (KDIGO) criteria, as recommended by the AHA.1 Qualifying metabolic risk factors included elevated fasting serum triglycerides (≥135 mg/dL), hypertension, diabetes, or metabolic syndrome (≥3 of the following: elevated waist circumference, low high density lipoprotein cholesterol (HDL) level [<40 mg/dL or <50 mg/dL for men or women, respectively], fasting serum triglycerides ≥150 mg/dL, elevated blood pressure [systolic blood pressure ≥130, diastolic blood pressure ≥80 mmHg, and/or use of blood pressure-lowering medications], or prediabetes). CKD stages were identified based on GFR and urinary albumin-to creatinine ratio.  **Stage 3: Subclinical CVD in CKM**  CKM Stage 3 was identified based of the presence of very-high-risk KDIGO CKD stages1 or a high-predicted 10-year CVD risk. 10-year cardiovascular risk was estimated with the AHA Predicting Risk of CVD EVENTs (PREVENT) equations.3 High risk was defined as ≥20% 10 year CVD risk (based on recommended thresholds). The PREVENT equations were developed and validated for adults 30-79 years of age. As such, risk was not estimated for adults <30 years. However, to minimize underestimation of CKD Stage 3, adults ≥80 years were not excluded from 10-year CVD risk. Instead, adults ≥80 years were assigned an age of 79 years when determining 10-year CVD risk to allow for conservative estimates. Further, PREVENT was developed for variables with the following ranges: total cholesterol 130-320 mg/dL, HDL 20-100 mg/dL, systolic blood pressure 90-200 mmHg, and GFR 14-140 mL/min/1.73m². To approximate PREVENT risk strata, values for these variables above or below these bounds were set to the upper or lower bounds of allowable values respectively (for example, total cholesterol of 330 mg/dL was set as 320 mg/dL). Cardiac biomarkers and cardiovascular imaging were not available to identify subclinical CVD.  **Stage 4: Clinical CVD in CKM**  CKM Stage 4 was identified based on self-reported established cardiovascular disease (coronary heart disease, angina, heart attack, heart failure, and stroke). Atrial fibrillation and peripheral artery disease were not included, as these data were not available. |

**Supplementary -table 2. Cox-regression analysis of risk factors for CVD mortality in individuals with CKM**

| Variables | Unadjusted | | Model 1 | | | Model 2 | | Model 3 | | |
| --- | --- | --- | --- | --- | --- | --- | --- | --- | --- | --- |
|  | HR (95% CI) | *P*-value | HR (95% CI) | *P* value | HR (95% CI) | | *P* value | | HR (95% CI) | *P* value |
| MASLD |  |  |  |  |  | |  | |  |  |
| No | ref |  | ref | ref | ref | | ref | | ref | ref |
| Yes | 2.66(1.90,3.73) | <0.0001 | 1.90(1.26, 2.86) | 0.002 | 1.76(1.18, 2.62) | | 0.01 | | 1.63(1.05, 2.52) | 0.03 |
| Age (years) |  |  |  |  |  | |  | |  |  |
| <65 | ref | ref | ref |  | ref | | ref | | ref | ref |
| ≥65 | 14.45(7.97,26.21) | <0.0001 | 13.91(7.19,26.91) | <0.0001 | 12.93(6.51,25.70) | | <0.0001 | | 8.90(4.65,17.02) | <0.0001 |
| Sex |  |  |  |  |  | |  | |  |  |
| Female | ref | ref | ref |  | ref | | ref | | ref | ref |
| Male | 1.62(1.08,2.44) | 0.02 | 1.79(1.24, 2.58) | 0.002 | 2.08(1.47, 2.93) | | <0.0001 | | 1.70(1.18, 2.45) | 0.004 |
| Ethnicity |  |  |  |  |  | |  | |  |  |
| Mexican American | ref | ref | ref | ref | ref | | ref | | ref | ref |
| Non-Hispanic Black | 1.51(0.81,2.79) | 0.19 | 1.44(0.78, 2.65) | 0.24 | 2.01(0.99, 4.10) | | 0.05 | | 1.70(0.83, 3.51) | 0.15 |
| Non-Hispanic White | 1.73(1.02,2.93) | 0.04 | 0.91(0.52, 1.60) | 0.75 | 1.64(0.79, 3.40) | | 0.18 | | 1.34(0.63, 2.84) | 0.45 |
| Other Hispanic | 0.87(0.44,1.69) | 0.67 | 0.77(0.41, 1.44) | 0.41 | 0.88(0.46, 1.70) | | 0.71 | | 0.84(0.42, 1.68) | 0.63 |
| Other Race - Including Multi-Racial | 1.01(0.35,2.90) | 0.98 | 0.76(0.26, 2.25) | 0.62 | 1.15(0.38, 3.54) | | 0.8 | | 1.03(0.34, 3.14) | 0.96 |
| BMI (kg/m2) |  |  |  |  |  | |  | |  |  |
| <30 | ref | ref | ref | ref | ref | | ref | | ref | ref |
| ≥30 | 1.08(0.72,1.62) | 0.72 | 0.86(0.54, 1.37) | 0.53 | 0.91(0.57, 1.45) | | 0.69 | | 0.92(0.57, 1.49) | 0.74 |
| Alcohol use |  |  |  |  |  | |  | |  |  |
| No |  |  |  |  | ref | | ref | | ref | ref |
| Yes | 0.62(0.39,1.00) | 0.05 |  |  | 0.76(0.47, 1.24) | | 0.28 | | 0.77(0.49, 1.20) | 0.25 |
| Smoke |  |  |  |  |  | |  | |  |  |
| No |  |  |  |  | ref | | ref | | ref | ref |
| Yes | 1.43(0.94,2.18) | 0.09 |  |  | 1.05(0.65, 1.67) | | 0.85 | | 0.94(0.58, 1.51) | 0.79 |
| Poverty (10000 dollar) (%) |  |  |  |  |  | |  | |  |  |
| ≤1 | ref | ref |  |  | ref | | ref | | ref | ref |
| 1.1-3 | 1.36(0.83,2.22) | 0.22 |  |  | 0.95(0.59, 1.53) | | 0.83 | | 0.97(0.61, 1.54) | 0.9 |
| >3 | 0.65(0.35,1.19) | 0.16 |  |  | 0.56(0.28, 1.10) | | 0.09 | | 0.62(0.31, 1.23) | 0.17 |
| Education (%) |  |  |  |  |  | |  | |  |  |
| High school or equivalent | ref | ref |  |  | ref | | ref | | ref | ref |
| Less than high school | 2.15(1.39,3.33) | <0.001 |  |  | 1.68(1.00, 2.84) | | 0.05 | | 1.60(0.94, 2.74) | 0.09 |
| College or above | 3.82(2.13,6.85) | <0.0001 |  |  | 2.41(1.07, 5.40) | | 0.03 | | 2.19(1.00, 4.79) | 0.05 |
| CVD |  |  |  |  |  | |  | |  |  |
| No | ref | ref |  |  |  | |  | | ref | ref |
| Yes | 10.47(6.94,15.79) | <0.0001 |  |  |  | |  | | 3.75(2.43, 5.77) | <0.0001 |

**Model 1** adjusted for baseline age (‘<65’ years, ‘≥65’ years), gender (‘Female’, ‘Male’), race (‘Mexican American’, ‘Non-Hispanic Black’, ‘Non-Hispanic White’, ‘Other Hispanic’, ‘Other Race - Including Multi-Racial’), BMI (<30kg/m^2^, ≥30 kg/m^2^); **Model 2** adjusted for covariates in model 1 plus smoke (‘Yes’ or ‘No’), alcohol use (‘Yes’ or ‘No’), education (‘College or above’, ‘High school or equivalent’, ‘Less than high school’), poverty (‘0-1’, ‘1.1-3’, ‘＞3’) . **Model 3** adjusted for covariates in model 2 plus CVD (‘Yes’ or ‘No’). MASLD, Metabolic Dysfunction-Associated Steatotic Liver Disease; CKM, Cardiovascular-kidney-Metabolic; HR, Hazard ratio; CI, Confidence interval; BMI, Body Mass Index; CVD, Cardiovascular disease; CKD, chronic kidney disease; DM, diabetes mellitus.

**Supplementary-table 3. The relationship between MASLD and CVD mortality in patients with CKM**

| Variables | Unadjusted |  | Model 1 |  | Model 2 |  | Model 3 |  |
| --- | --- | --- | --- | --- | --- | --- | --- | --- |
|  | HR (95%CI) | *P* value | HR (95%CI) | *P* value | HR (95%CI) | *P* value | HR (95%CI) | *P* value |
| Non-MASLD | ref |  | ref |  | ref |  | ref |  |
| MASLD | 2.66(1.90,3.73) | <0.0001 | 1.90(1.26, 2.86) | 0.002 | 1.76(1.18, 2.62) | 0.01 | 1.63(1.05, 2.52) | 0.03 |

**Model 1** adjusted for baseline age (‘<65’ years, ‘≥65’ years), gender (‘Female’, ‘Male’), race (‘Mexican American’, ‘Non-Hispanic Black’, ‘Non-Hispanic White’, ‘Other Hispanic’, ‘Other Race - Including Multi-Racial’), BMI (<30kg/m^2^, ≥30 kg/m^2^); **Model 2** adjusted for covariates in model 1 plus smoke (‘Yes’ or ‘No’), alcohol use (‘Yes’ or ‘No’), education (‘College or above’, ‘High school or equivalent’, ‘Less than high school’), poverty (‘0-1’, ‘1.1-3’, ‘＞3’) . **Model 3** adjusted for covariates in model 2 plus CVD (‘Yes’ or ‘No’). MASLD, Metabolic Dysfunction-Associated Steatotic Liver Disease; CKM, Cardiovascular-kidney-Metabolic; HR, Hazard ratio; CI, Confidence interval; BMI, Body Mass Index; CVD, Cardiovascular disease; CKD, chronic kidney disease; DM, diabetes mellitus.

**Supplementary-table 4. Stratified analysis of MASLD and the risk of CVD mortality in individuals with CKM**

| Variable | HR（95% CI） | | | *P* for interaction |
| --- | --- | --- | --- | --- |
|  | Non-MASLD | MASLD | *P* for trend |  |
| Age (years) |  |  |  | 0.04 |
| ≤65 | ref | 1.80(1.08,3.00) | 0.02 |  |
| >65 | ref | 1.07(0.86,1.33) | 0.54 |  |
| Gender |  |  |  | 0.76 |
| Male | ref | 1.44(1.06,1.96) | 0.02 |  |
| Female | ref | 1.00(0.70, 1.44) | 0.99 |  |
| BMI (kg/m2) |  |  |  | 0.12 |
| ≤30 | ref | 0.62(0.39, 0.98) | 0.04 |  |
| ＞30 | ref | 0.84(0.66,1.07) | 0.15 |  |
| Ethnicity |  |  |  | < 0.001 |
| Mexican American | ref | 1.55(0.84, 2.88) |  |  |
| Non-Hispanic Black | ref | 17.70(1.90,165.01) |  |  |
| Non-Hispanic White | ref | 0.43(0.13, 1.48) |  |  |
| Other Hispanic | ref | 3.00( 0.60, 15.03) |  |  |
| Other Race - Including Multi-Racial | ref | 6.70(0.60, 74.25) |  |  |
| **Hypertension** |  |  |  | 0.27 |
| No | ref | 1.23(0.77, 1.97) | 0.38 |  |
| Yes | ref | 1.15(0.87,1.53) | 0.32 |  |
| DM |  |  |  | 0.01 |
| No | ref | 0.76(0.55, 1.04) | 0.08 |  |
| Yes | ref | 1.14(0.85,1.53) | 0.39 |  |
| CKD |  |  |  | 0.08 |
| No | ref | 1.36(0.94,1.98) | 0.1 |  |
| Yes | ref | 0.96(0.72,1.29) | 0.81 |  |
| CVD |  |  |  | 0.43 |
| No | ref | 1.26(0.91,1.73) | 0.17 |  |
| Yes | ref | 1.21(0.81,1.79) | 0.36 |  |
| CKM Syndrome |  |  |  | < 0.001 |
| Stage 1 | ref | 0.16( 0.03, 0.87) | 0.03 |  |
| Stage 2 | ref | 0.22(0.09, 0.52) | <0.001 |  |
| Stage 3 | ref | 1.41(0.57, 3.50) | 0.46 |  |
| Stage 4 | ref | 0.97(0.57, 1.63) | 0.9 |  |

adjusted for baseline age (‘<65’ years, ‘≥65’ years), gender (‘Female’, ‘Male’), ethnicity (‘Mexican American’, ‘Non-Hispanic Black’, ‘Non-Hispanic White’, ‘Other Hispanic’, ‘Other Race - Including Multi-Racial’), BMI (<30kg/m^2^, ≥30 kg/m^2^), Hypertension (‘Yes’ or ‘No’), DM (‘Yes’ or ‘No’), CKD (‘Yes’ or ‘No’), CVD (‘Yes’ or ‘No’), CKM Syndrome (‘Stage 1’, ‘Stage 2’, ‘Stage 3’, ‘Stage 4’). MASLD, Metabolic Dysfunction-Associated Steatotic Liver Disease; HR, Hazard ratio; CI, Confidence interval; BMI, Body Mass Index; CKD, chronic kidney disease; DM, diabetes mellitus; CVD, Cardiovascular disease; CKM, Cardiovascular-kidney-Metabolic.

**Supplementary-table 5 The relationship between MASLD and CVD mortality in patients with CKM after deleting missing values**

| Variables | Unadjusted |  | Model 1 |  | Model 2 |  | Model 3 |  |
| --- | --- | --- | --- | --- | --- | --- | --- | --- |
|  | HR (95%CI) | *P* value | HR (95%CI) | *P* value | HR (95%CI) | *P* value | HR (95%CI) | *P* value |
| Non-MASLD | ref |  | ref |  | ref |  | ref |  |
| MASLD | 2.66(1.90,3.72) | <0.0001 | 2.00 (1.33, 3.01) | <0.001 | 2.00(1.30, 3.08) | 0.002 | 1.84(1.14, 2.96) | 0.012 |

**Model 1*^a^*** adjusted for baseline age (‘<65’ years, ‘≥65’ years), gender (‘Female’, ‘Male’), ethnicity (‘Mexican American’, ‘Non-Hispanic Black’, ‘Non-Hispanic White’, ‘Other Hispanic’, ‘Other Race - Including Multi-Racial’), BMI (<30kg/m^2^, ≥30 kg/m^2^); **Model 2*^b^*** adjusted for covariates in model 1 plus smoke (‘Yes’ or ‘No’), alcohol use (‘Yes’ or ‘No’), education (‘College or above’, ‘High school or equivalent’, ‘Less than high school’), poverty (‘0-1’, ‘1.1-3’, ‘＞3’); **Model 3*^c^*** adjusted for covariates in model 2 plus CVD (‘Yes’ or ‘No’). MASLD, Metabolic Dysfunction-Associated Steatotic Liver Disease; CKM, Cardiovascular-kidney-Metabolic; HR, Hazard ratio; CI, Confidence interval; BMI, Body Mass Index; CVD, Cardiovascular disease.

**Supplementary-table 6. The relationship between MASLD and CVD mortality in patients with CKM after deleting missing values(换成FTI诊断标准)**

| Variables | Unadjusted |  | Model 1 |  | Model 2 |  | Model 3 |  |
| --- | --- | --- | --- | --- | --- | --- | --- | --- |
|  | HR (95%CI) | *P* value | HR (95%CI) | *P* value | HR (95%CI) | *P* value | HR (95%CI) | *P* value |
| Non-MASLD | ref |  | ref |  | ref |  | ref |  |
| MASLD | 1.958(1.322,2.899) | <0.001 | 1.820(1.158,2.860) | 0.009 | 1.749(1.115,2.741) | 0.015 | 1.676(1.068,2.630) | 0.025 |

**Model 1*^a^*** adjusted for baseline age (‘<65’ years, ‘≥65’ years), gender (‘Female’, ‘Male’), ethnicity (‘Mexican American’, ‘Non-Hispanic Black’, ‘Non-Hispanic White’, ‘Other Hispanic’, ‘Other Race - Including Multi-Racial’), BMI (<30kg/m^2^, ≥30 kg/m^2^); **Model 2*^b^*** adjusted for covariates in model 1 plus smoke (‘Yes’ or ‘No’), alcohol use (‘Yes’ or ‘No’), education (‘College or above’, ‘High school or equivalent’, ‘Less than high school’), poverty (‘0-1’, ‘1.1-3’, ‘＞3’); **Model 3*^c^*** adjusted for covariates in model 2 plus CVD (‘Yes’ or ‘No’). MASLD, Metabolic Dysfunction-Associated Steatotic Liver Disease; CKM, Cardiovascular-kidney-Metabolic; HR, Hazard ratio; CI, Confidence interval; BMI, Body Mass Index; CVD, Cardiovascular disease.
